# Determinants of Choice of Abortion Methods among Women in Ghana; Does Knowledge on legality of Abortion Matter?

**DOI:** 10.1101/2020.07.02.20144923

**Authors:** Desmond Klu, Donatus Yaw Atiglo, Alfred Kwesi Manyeh, Mustapha Immurana, Maxwell Dalaba

**Author notes:** Corresponding author Desmond Klu (PhD). Co-authors.

## Abstract

**Objective:** This study therefore examines the role knowledge on legal abortion and other factors play in the choice of abortion methods among women within a ten-year period.

**Design:** The study use data from the 2007 and 2017 Ghana Maternal Health Surveys. A total sample of 2,432 women aged 15-49 whose activities related to their most recent induced abortion were selected. This study, however, limits the analysis to a subpopulation of women who have terminated a pregnancy between 2002 to 2007 and 2012 to 2017. The made use of binary logistic regression analysis to show the relationship between choice of abortion methods and knowledge of legality of abortion.

**Setting:** Ghana

**Participants:** Women who had practice induce abortion

**Main outcome measure:** Choice of Abortion methods

**Results:** Likelihood of unsafe abortion practices was high among women who had knowledge on legality of abortion, women with no formal education, single women, rural women and women whose partners did not know about the pregnancy.

**Conclusion:** Increasing knowledge of legal status on abortion among women corresponded with an increased in the use of unsafe abortion methods over the decade has implication for poor reproductive health outcome and increased maternal mortality among women in Ghana.

**Article summary:** Strengths and limitations of the study

► Use of two rounds of nationally representative data to show knowledge of women on legality abortion that can be generalized.
► Measurement of Safe and Unsafe Abortion methods which involved combination safe and safe provider, place of abortion and methods of abortion which reflect a robust measurement of abortion.
► Interesting outcome of high unsafe abortion practices among women who have good knowledge on the legality of abortion under some circumstances
► The use of secondary data prevented us from including other important variables in our study to was one major limitation of our study
► Another limitation includes the inability to explore further using qualitative methods that will provide detailed explanation to the reasons of high unsafe abortion practices among highly knowledgeable women of legality of abortion

## Background

Globally, it is estimated annually that about 56 million induced abortions occur^1^, which about 55 percent use unsafe methods. Within the African region, it is estimated that close to 8.2 million women commit abortion.^2^ For instance, 34 per 1000 women in South, Middle and Eastern Africa and 31 per 1000 women in West Africa are estimated to commit abortion.^3^ Globally, the use of unsafe abortion method is more prevalent in settings where access to legal abortion services is highly controlled.^4^ Unsafe abortion therefore contributes to 7.9 percent of maternal related deaths and hence a major cause of morbidity in women.^5^

Ghana has a liberal abortion law since 1985.^6^ In that, abortion may be allowed under some circumstances such as rape, incest, and when life of the mother or unborn child is at risk.^7^ Despite the liberalisation in the law, use of unsafe abortion methods increased from 45 percent in 2007 to 62 percent in 2017.^7 8^ There have also been efforts by government of Ghana to reduce the practise of unsafe abortion and as a strategy to reduce maternal mortality. This includes but not limited to the introduction of Reducing Maternal Mortality and Morbidity (R3M) programmme, which aimed at not only improving access to family planning services but also comprehensive abortion care services.^7 9^ There have also been high supply and availability of modern abortion medication such as mifepristone and misoprostol on the Ghanaian market.^10 11^ These efforts however, had barely impacted on the reduction of the prevalence of unsafe abortion methods in Ghana.^12^

The availability of abortion related services, programmes and policies and the impact of these interventions may be influenced by some individual, household, community, socio-cultural, political, economic and systemic factors.^13-16^ Therefore, a vivid comprehension of abortion service provision and the choice of abortion methods (safe and unsafe) are paramount, especially in the context of negative perceptions and misconception surrounding abortion. Moreover, in recent times, there is high recognition of the extent to which the choice of an abortion method implicates the sexual and reproductive life of women and the health care system.^17^

Empirically, a large body of literature have identified the determinants of choice of safe methods.^6 9 19 20^ and unsafe abortion methods.^21-25^ Other studies also examine the association between women’s knowledge of abortion law and the practice of induced abortion.^26-29^ Further, while some studies use hospital-based data in examining this phenomenon ^27 30-32^ and community-based surveys^26 28 33 34^, others use nationally representative surveys. ^20 25 35 36^ These previous studies did not fully explore and examine how demographic, socio-economic factors influence women’s knowledge on the legality of abortion under some conditions (rape, incest, risk to mothers and foetus health) and how that interaction determines their choice of abortion methods. Again, no study, to the best of my knowledge has examined how factors affecting the choice of abortion method and knowledge on legality of abortion change over time using nationally representative data.

The present study seeks to examine how women’s knowledge on legal status of abortion, demographic and socio-economic factors influence the choice of abortion in Ghana using the 2007 and 2017 Ghana Maternal Health Survey. Thus, the study explores and examines how women’s demographic, socio-economic characteristics have shaped their knowledge on the legality of abortion under some circumstance and how that affect their choice of abortion methods over a decade (2007-2017).

### Brief overview of women’s knowledge on legality of abortion and abortion practices

Towards the end of the 20^th^ century, the issue of abortion was considered in many countries as illegal with moral and religious connotations. It was also highly associated with health complications and deaths especially, when it is unsafely done.^37^

However, in recent times, there has been liberalization of laws on abortion both in the developed and developing world and Ghana is not an exception. These liberalizations came along with promotion and advocacy of modern contraceptive use to ensure the reduction in unsafe abortion rate and its corresponding mortality.^37 38^

In 1985, Ghana witnessed the repeal of the 1960 Criminal Code (Act 29), which gave birth to the Provisional National Defense Council (PNDC) Law 102 liberalising law on abortion in Ghana.^39 40^ The new law states that, induced abortion will only be accepted as legal under the following circumstances

a. If conception results from rape (non-consensual penetrative sexual intercourse)
b. Defilement of a female minor (consensual or non-consensual sex with a girl below the age of 16/ mentally disabled)
c. Incest ((consensual or non-consensual sex with a female of blood relation)
d. Pregnancy that is considered a threat to the health/life of the women or foetus

Studies have argued that these legal amendments and ramifications to Ghana’s abortion law is open to many interpretations because of the wide application of social, mental and physical health used as a basis for practicing induced abortion.^8 19^ These studies therefore suggested that there should be more liberalisation and amendments to the abortion laws. This will empower women and enhance their right and ability to make an informed choice when it comes to abortion decision process. Meanwhile, the question is ‘How well are women knowledgeable of the legalities surrounding the issue of abortion?’ ‘Even if they are well-informed on the legalities of abortion, which factors (internal and external) can affect their choice of an abortion method?

Studies have attempted to provide possible explanation to the above questions. For instance, a study by Gbagbo ^27^ in Ghana found that less than half of women had knowledge on legality of abortion under some conditions. The study further found that this knowledge on legality of abortion was among women who were highly educated, and it influenced their decision to seek safe abortion services. Again, studies have indicated that women who knew abortion was legal under some circumstances were more likely to choose safe abortion methods. ^27 41 42^

Also, some studies argue that in an environment where strict cultural systems, moral and religious laws and conventions prohibits abortion practices, knowledge on legality of abortion can be influenced by several factors at the individual, household, community, societal and national levels.^16 20 22 23^ These factors include but not limited to age ^43^, marital status ^33^, ethnicity ^44^, geographical location ^45^ and economic circumstances. ^46^ The interaction among these multidimensional factors does influence knowledge on legality of abortion and the decision-making process on the choice of abortion methods.^18^

It is clear from the literature that the relationship between knowledge on legality of abortion under some conditions and abortion is mediated by factors at the micro and macro levels. However, what literature has been silent on is how the changes in the micro and macro level factors might affect the relationship between knowledge of abortion legality and choice of abortion method. This is the literature gap that this study seeks to fill by examining how the changes in these factors affect legality knowledge of abortion and abortion decision making nexus.

## Methods and Materials

The study use data from the 2007 and 2017 Ghana Maternal Health Surveys (GMHSs). The 2007 and 2017 GMHSs were spearheaded by the Ghana Statistical Service with technical support from Inner City Fund (ICF) Macro International through the DHS Program, funded by the United States Agency for International Development (USAID), Government of Ghana, the European Union (EU) and the United Nations Population Fund (UNFPA). The sampling frame adopted was from the 2000 and 2010 Ghana Population and Housing Censuses (PHCs). The GMHSs used a multistage stratified cluster sampling method to select the eligible enumeration areas and households. Further details of the survey methodological and sampling procedures and questionnaires used can be accessed in the final report. ^47 48^ Women of reproductive age 15-49 years who were permanently resident in selected household a night before the survey were eligible to be interviewed.

The study made use of the women’s data file and for the two rounds of GHMS conducted in 2007 and 2017. This study, however, limits the analysis to a subpopulation of women who have terminated a pregnancy between 2002 to 2007 and 2012 to 2017 due to the dependent variable of interest (choice of abortion methods). A total sample of 2,432 (552 in 2007 and 1,880 in 2017) women whose activities related to their most recent induced abortion were selected.

### Variables

The outcome variable is choice of abortion method. It was categorized in a binary form as “0” for “safe” and “1” for unsafe from the variable ‘type of method, provider and place of abortion’. The ‘safe’ abortion methods came from medical method such as misoprostol (Cytotec tablets), mifepristone, dilation and curettage (D&C), dilation and evacuation (D&E), vacuum aspiration, saline installation, catheter and other medical injections. In terms of safe provider, it was defined as seeking abortion services from a medical doctor, nurse/midwife-these are legally certified health professionals mandated to provide these services. The use of ‘safe’ place for abortion was defined as seeking abortion services from a government/private hospital/ health centre because of the requisite support equipment and hygiene is likely to be present in these facilities Any woman who responded in the affirmative to all these three measures (safe methods, provider and place) in terminating her pregnancy was classified to have used safe abortion method. However, if she used only one or two of these measures (using D&C, performed by a doctor but in an unapproved setting) is considered as ‘unsafe’. The ‘unsafe’ method on the other hand, was operationally defined as the termination of pregnancy using non-medically certified methods. These methods include drinking milk/coffee, drinking herbal concoction, drinking other home remedies, use of herbal enema, inserting herb/object into the vagina, excessive physical activity, taking of unknown tablets and heavy massage. Unsafe providers in this context include pharmacist/chemical seller, traditional birth attendant, relative/friend and community health workers. With respect to unsafe place of abortion, we classified them as women receiving induced abortion services at respondent’s own home, home of traditional birth attendants, friend/relative’s home or pharmacy. Any woman who affirmed to have used any of these methods, providers or place to terminate pregnancy was classified to have used unsafe abortion method.

The main explanatory variable used was knowledge on legality of abortion. The survey, women who had ever had an induced abortion were asked if they thought induced abortion is legal in Ghana. Those who answered in affirmative were further asked under what circumstances it is legal. Therefore, the proportion who stated they know induced abortion is legal in Ghana comprises those who have heard of abortion and stated rightly the circumstances under which induced abortion can be performed. The other explanatory variables are age, religious affiliation of the women, educational level, ecological zones, place of residence, ethnicity and parity. Table 1 illustrates the categorization of each variable (see Appendix 1).

**Table 1:** Categorisation of Variables.

| *Variable* | *Categorisation* |
| --- | --- |
| Age | 15-29<br>30-39<br>40-49 |
| Religious Affiliation | Catholic<br>Protestant<br>Moslem<br>Pentecostal/Charismatic<br>No religion |
| Educational Level | No education<br>Primary<br>Secondary+ |
| Ethnicity | Akan<br>Ga/Dangme<br>Ewe<br>Mole Dagbani |
| Marital Status | Married<br>Cohabiting<br>Not Married |
| Parity | 0<br>1-2<br>3+ |
| Ecological Zones | Coastal Zone (Western, Central, Greater Accra & Volta)<br>Middle-Belt Zone (Eastern, Ashanti & Brong Ahafo)<br>Northern Zone (Northern, Upper East & Upper West) |
| Place of Residence | Urban<br>Rural |
| Partners' attitude toward Abortion | Favored<br>Opposed<br>Neutral<br>Didn't know about pregnancy |

### Statistical Analysis

The analytical strategy comprises both descriptive (frequencies) and inferential (chi-square & binary logistic) statistical analyses. Thus, the association between the predictor variables and the outcome variables were examined in bivariate (chi square) and multivariate logistic regression analysts. In the analysis, the odd ratios with 95% confidence interval was calculated and statistical significance was set at a p-value of <0.05. All analyses were done using SPSS V.20.

### Ethics

The ICF Institutional Review Board (IRB) approved the protocol for the 2007 and 2017 GMHSs. However, ethnical approval was not needed for this study since it involved secondary data analysis devoid of personal identifies to women and their households. Nonetheless, the researcher obtained permission from ICF for the use of the datasets and the terms of use have been strictly adhered to.

## Results

The results on background characteristics of women who induced abortion, the prevalence of women’s knowledge on legality of abortion and abortion methods is presented in Table 2 (see appendix 2). It shows that in the two surveys, majority of women in Ghana do not know that abortion is legal under some condition relative to those who have some knowledge. However, there has been an increase in the proportion of women who knows about the legality of abortion under some conditions in Ghana from 6.2% in 2007 to 10.7% in 2017.

**Table 2:** Background characteristics, prevalence of women’s knowledge on legality of abortion and methods.

| *Variable* | *2007* |  | *2017* |  |
| --- | --- | --- | --- | --- |
| **Knowledge on Legality of Abortion** | **Frequency** | **Percent** | **Frequency** | **Percent** |
| Yes | 34 | 6.2 | 200 | 10.7 |
| No | 518 | 93.8 | 1680 | 89.3 |
| **Abortion Methods** |  |  |  |  |
| Safe Methods | 362 | 65.6 | 1402 | 74.6 |
| Unsafe Methods | 190 | 34.4 | 478 | 25.4 |
| **Current Age** |  |  |  |  |
| 15-29 | 385 | 68.8 | 1242 | 66.1 |
| 30-39 | 135 | 24.5 | 521 | 27.7 |
| 40-49 | 37 | 6.7 | 117 | 6.2 |
| **Religious Affiliation** |  |  |  |  |
| Catholic | 57 | 10.4 | 163 | 8.7 |
| Protestant | 219 | 39.7 | 498 | 26.5 |
| Moslem | 29 | 5.3 | 127 | 6.8 |
| Pentecostal/Charismatic | 228 | 41.2 | 1044 | 55.5 |
| No religion | 19 | 3.4 | 48 | 2.5 |
| **Educational Level** |  |  |  |  |
| No Education | 42 | 7.7 | 162 | 8.6 |
| Primary | 131 | 23.8 | 298 | 15.9 |
| Secondary+ | 379 | 68.6 | 1420 | 75.5 |
| **Ethnicity** |  |  |  |  |
| Akan | 337 | 61.0 | 1133 | 60.3 |
| Ga/Dangme | 67 | 12.0 | 171 | 9.1 |
| Ewe | 96 | 17.4 | 335 | 17.8 |
| Mole-Dagbani | 53 | 9.5 | 242 | 12.9 |
| **Marital Status** |  |  |  |  |
| Married | 226 | 40.9 | 438 | 23.3 |
| Cohabiting | 89 | 16.2 | 682 | 36.3 |
| Not Married | 237 | 42.9 | 761 | 40.5 |
| **Parity** |  |  |  |  |
| 0 | 250 | 45.3 | 534 | 28.4 |
| 1-2 | 238 | 43.1 | 861 | 45.8 |
| 3+ | 64 | 11.6 | 458 | 25.8 |
| **Ecological Zones** |  |  |  |  |
| Coastal Zone | 251 | 45.4 | 1032 | 54.9 |
| Middle Belt | 290 | 52.4 | 795 | 42.3 |
| Northern Zone | 13 | 2.2 | 53 | 2.8 |
| **Place of residence** |  |  |  |  |
| Urban | 330 | 59.8 | 1230 | 65.4 |
| Rural | 222 | 40.2 | 650 | 34.6 |
| **Partners' attitude toward Abortion** |  |  |  |  |
| Favored | 326 | 59.0 | 992 | 52.8 |
| Opposed | 102 | 18.5 | 337 | 17.9 |
| Neutral | 24 | 4.4 | 160 | 8.5 |
| Didn't know about pregnancy | 100 | 18.1 | 391 | 20.8 |

Source: Computed from Ghana Maternal Health Surveys (GMHS) 2007 & 2017

Regarding choice of abortion methods, majority of the women reported using safe methods to abort their babies compared to those who used the unsafe methods. It is also important to note that the proportion of women who used unsafe method have decreased over the decade which increased from 34.4% in 2007 to 25.4% in 2017.

Concerning respondents’ background characteristics, Table 2 indicates that in the two surveys, relatively higher proportion of women practicing induced abortion was within the age group 15-29 years: 68.8% in 2007, 66.1% in 2017 compared to other age categories. With respect to religious affiliation, in 2007, the highest proportion of women who aborted their pregnancies were Pentecostal/Charismatic (41.2%) whilst the least were women who had no religious affiliations (3.4%). Indeed, a decade later, the practice of abortion was still high among women who belong to the Pentecostal/Charismatic faith (55.5%). Women with secondary or higher level of education constitute the highest proportion of those who had ever had an abortion in the two surveys (68.6%: 2007; 75.5%: 2017). With regards to ethnicity, the results clearly show high rate of abortion among Akan women compared to the other ethnic groups in 2007 and 2017.

Concerning marital status, the result shows higher proportion of single women (42.9%) aborting their pregnancies in 2007 compared to those cohabiting (16.2%) and married (40.9%). Again, in 2017, higher incidence of abortion was recorded among women who are not married (40.5%) relative to the married (23.3%) and cohabiting ones (36.3%). In terms of parity, the prevalence of abortion was higher among women who had no children (45.3%) relative women with one to two children (43.1%) and three or more children (11.6%) in 2007. The observation looks different in 2017, where higher proportion of women who committed abortion were those with one to two children (54.9%) compared to those who had none (28.4%) and three or more children (25.8%).

In terms of the ecological zones respondents’ dwell, the result vividly shows the practice of abortion was high among women who dwell in the middle belt zone relative to the other zones in 2007. However, in 2017, the rate of abortion was highest among women who reside in the coastal zones compared to the other zones. It was observed that the practice of induced abortions was more common among women in the urban areas within the decade.

In addition, rate of induced abortion was high among women whose partners were in favour of the abortion in the two survey years compared to partners opposed it, were neutral and did not know about the pregnancy. For instance, 59 percent of women whose partners showed a favourable attitude towards abortion committed an abortion in 2007 as against 18.5 percent whose partners opposed abortion, who were indifferent (4.4%) towards it and who were not aware of the pregnancy (18.1%). Similar observation was made in 2017, where about 53 percent of women aborted their pregnancies because their partners were in favour of it. However, the proportion reduced to about 18 percent for partners who opposed the abortion, who developed a neutral (8.5%) attitude towards it and not aware of the pregnancy had a proportion of about 21 percent.

### Association between women’s knowledge on abortion legality, background characteristics and choice of abortion methods

This section examined the association between women’s knowledge on abortion legality, choice of abortion methods and background characteristics. These are presented in Table 3(see appendix 3). The practice of unsafe abortion was higher among women, who had no knowledge on the legality of abortion under some circumstances in 2017 and women within the age group 15-29 years in 2007. Regarding educational level and abortion methods, the result shows, unsafe abortion practices were more common among women with primary level relative to those with no education and secondary/higher education in 2007. However, a decade later, the use of unsafe abortion method was high among women with no education. With ethnicity, women who belong to Ewe ethnic group recorded a high rate of unsafe abortion. The practice of unsafe abortions was most common among women who were cohabiting and dwelling in the rural areas. With regards to parity, the proportion using unsafe methods were recorded among women with no children in 2007 but in 2017, it was found more common among women with 5 or more children. In both surveys, the prevalence of unsafe induced abortion was high among women whose partners had no knowledge about their pregnancies compared to partners who were either opposed to it, favoured it or indifferent about the decision to abort.

**Table 3:**
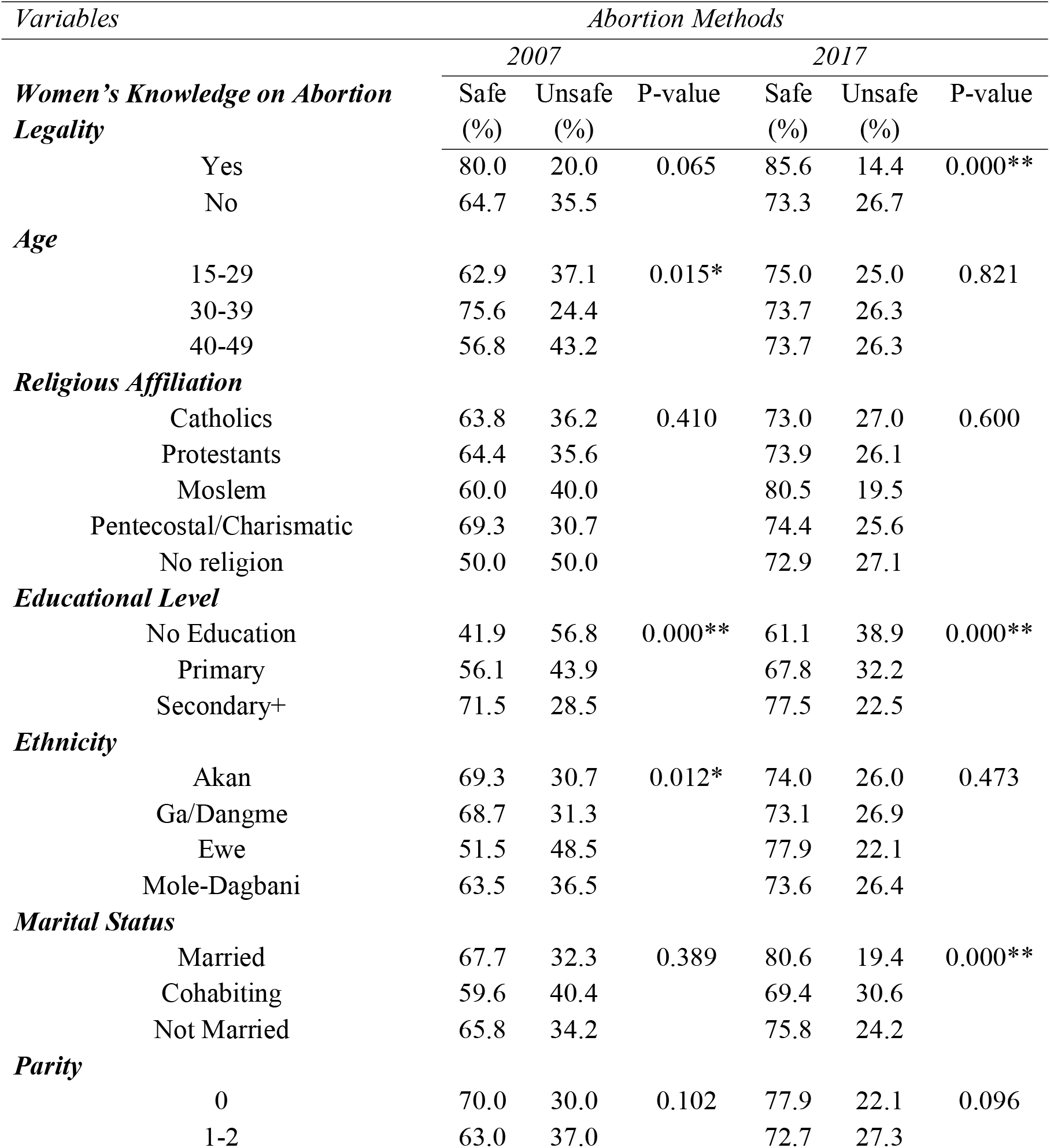
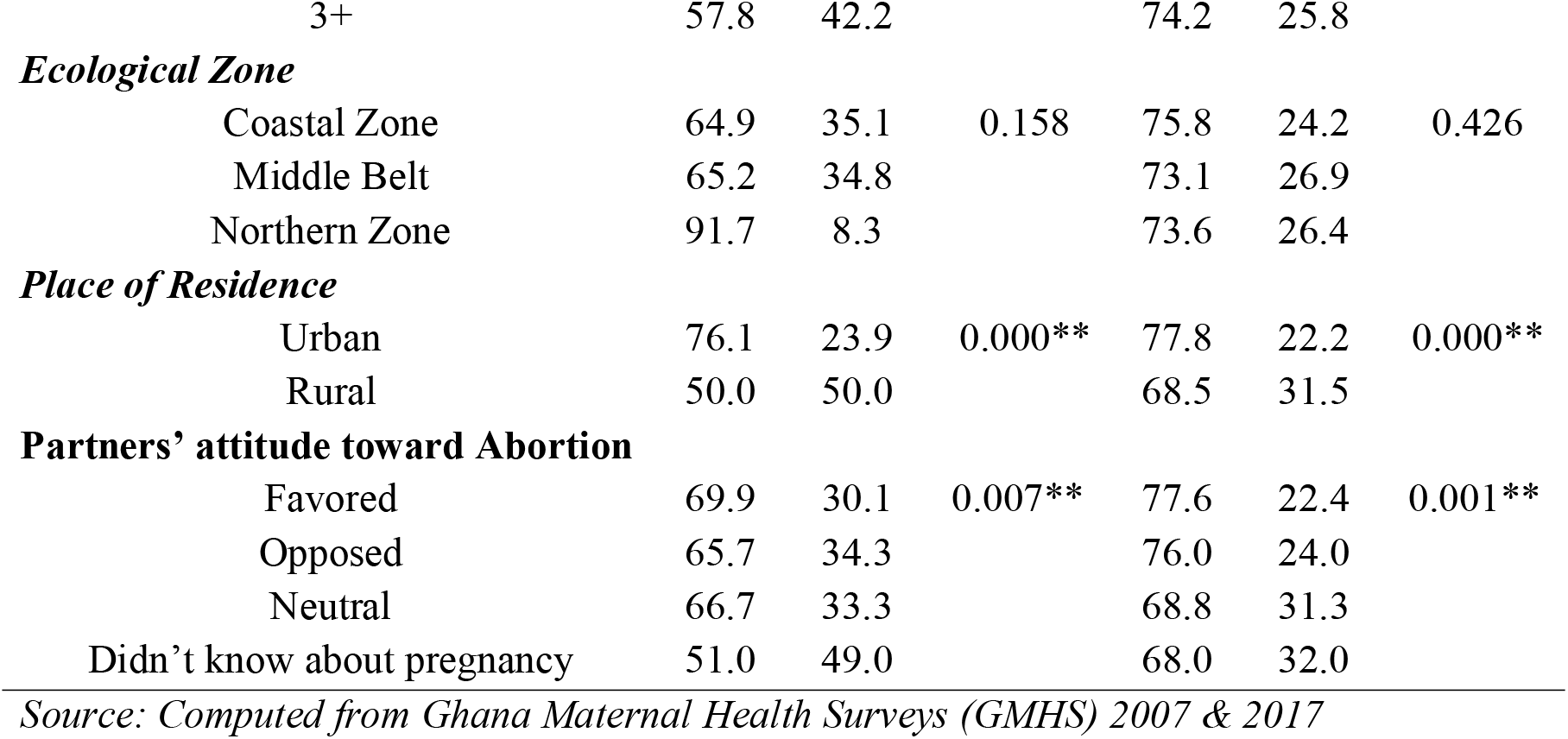
Association between women’s knowledge on abortion legality, background characteristics and choice of abortion methods.

### Predictors of Choice of Abortion Methods

Table 4 (see appendix 4) presents the results of the binary logistic regression analyses of the likelihood of women adopting safe or unsafe abortion methods in Ghana taking into consideration their knowledge on the legality of abortion under some condition as well as their background characteristics. From the results, women who knew the legal status of abortion in Ghana (OR=0.53, 95% CI: 0.35-0.80) were less likely to choose unsafe abortion method and women who are 15-29 years old (OR=10.29, 95%CI: 5.52-19.17). Women with secondary and higher education (OR=0.47, 95% CI: 0.32-0.68), married women and women residing in urban areas were all less likely to choose unsafe methods of abortion. The analysis further shows that, women with either one or two children (OR=2.22; 95%CI: 1.19-4.15) within the decade are more likely to practice abortion using unsafe methods.

**Table 4:** Binary Logistic Regression of the predictors of choice of abortion methods.

| Independent Variables | 2007 |  | 2017 |  |
| --- | --- | --- | --- | --- |
|  | OR | (95%CI) | OR | (95%CI) |
| **Women's Knowledge on Abortion** |  |  |  |  |
| **Legality** |  |  |  |  |
| Yes | 1.60 | (0.61-4.16) | 1.85*** | (1.21-2.82) |
| No | Ref |  | Ref |  |
| **Age** |  |  |  |  |
| 15-29 | 0.82 | (0.33-2.02) | 0.91 | (0.55-1.49) |
| 30-39 | 0.27** | (0.11-0.68) | 1.03 | (0.64-1.67) |
| 40-49 | Ref |  | Ref |  |
| **Religious Affiliation** |  |  |  |  |
| Catholic | 0.79 | (0.24-2.54) | 1.24 | (0.58-2.66) |
| Protestants | 0.96 | (0.33-2.78) | 1.24 | (0.61-2.52) |
| Moslem | 2.39 | (0.51-11.24) | 0.71 | (0.30-1.64) |
| Pentecostal/Charismatic | 0.77 | (0.27-2.25) | 1.19 | (0.60-2.38) |
| No religion | Ref |  | Ref |  |
| **Educational Level** |  |  |  |  |
| No Education | Ref |  | Ref |  |
| Primary | 0.50 | (0.23-1.10) | 0.69 | (0.45-1.05) |
| Secondary+ | 0.40* | (0.19-0.84) | 0.44*** | (0.30-0.64) |
| **Ethnicity** |  |  |  |  |
| Akan | 0.95 | (0.41-2.23) | 0.89 | (0.59-1.34) |
| Ga/Dangme | 0.68 | (0.25-1.81) | 0.97 | (0.57-1.66) |
| Ewe | 1.84 | (0.73-4.67) | 0.66 | (0.41-1.05) |
| Mole-Dagbani | *Ref* |  | *Ref* |  |
| **Marital Status** |  |  |  |  |
| Married | 0.80 | (0.49-1.31) | 0.70* | (0.51-0.97) |
| Cohabiting | 0.91 | (0.51-1.61) | 1.21 | (0.94-1.57) |
| Not Married | *Ref* |  | *Ref* |  |
| **Parity** |  |  |  |  |
| 0 | *Ref* |  | *Ref* |  |
| 1-2 | 1.34 | (0.84-2.14) | 1.17 | (0.88-1.56) |
| 3+ | 2.14 | (0.97-4.73) | 0.91 | (0.62-1.34) |
| **Ecological Zone** |  |  |  |  |
| Coastal Zone | *Ref* |  | *Ref* |  |
| Middle Belt | 0.92 | (0.59-1.42) | 1.06 | (0.84-1.34) |
| Northern Zone | 0.10* | (0.01-0.98) | 0.99 | (0.48-2.06) |
| **Place of Residence** |  |  |  |  |
| Urban | 0.29*** | (0.19-0.44) | 0.68*** | (0.54-0.85) |
| Rural | *Ref* |  | *Ref* |  |
| **Partners' attitude toward Abortion** |  |  |  |  |
| Favored | 0.38*** | (0.23-0.65) | 0.62*** | (0.47-0.81) |
| Opposed | 0.57 | (0.30-1.09) | 0.67* | (0.48-0.95) |
| Neutral | 0.50 | (0.18-1.41) | 0.92 | (0.61-1.39) |
| Didn't know about pregnancy | *Ref* |  | *Ref* |  |

Source: Computed from Ghana Maternal Health Surveys (GMHS) 2007 & 2017

In sum, between 2007 and 2017, women’s knowledge on legal status of abortion, age, education, marital status, parity and place of residence were all significant predictors of choice of abortion methods.

## Discussion

This study was designed to examine the influence of women’s knowledge on legal status of abortion, demographic and socio-economic factors on the choice of abortion in Ghana using the 2007 and 2017 Ghana Maternal Health Surveys. The findings illustrate that there has been an increase in the proportion of women practicing unsafe abortions from 12.6% in 2007 to 25.4% in 2017. This means the prevalence of unsafe abortions have doubled over the decade among women in Ghana. Similar findings were of high rate of abortion incidence were found by other studies recently in Ghana ^20 25^, Ethiopia ^49^ and Nepal. ^50^

In terms of the demographic and socio-economic influencers of abortion, the findings showed that younger women (15-29 years) relative to older women were more likely to choose unsafe methods of abortion. This finding is like findings of earlier studies from Ghana ^40^ and other countries.^50 55^ The possible explanation to this occurrence is that young women tend to practice more risky sexual behaviours, are likely to be victims of coerced sexual intercourse, and rape that can lead to unintended pregnancies ^12 24^ and lack financial resources to take care of the pregnancy. ^54^ Therefore, the decision to go for abortion and the method to use coupled with the absence of financial, emotional and social support compel these young women to choose the unsafe methods which are easily accessible and relatively cheaper.

With regards to education, the findings of the study showed that women with secondary/higher education are significantly less likely to use unsafe abortion methods compared to women with no education. This outcome supports the finding of other studies in Ghana ^6 20^ and in Bangladesh. ^56^ The plausible explanation to this is the greater access to knowledge and financial resources which enable them to have access to safer abortion methods and be well informed on the consequences of unsafe abortion methods.

In terms of marital status and the choice of abortion methods, the results show that married women are significantly less likely to use unsafe method during abortion compared to women who are cohabiting and unmarried. This finding is in concordance with findings of other studies. ^33 57^ It is strongly believed that married women who are in stable unions are likely to receive financial, social and psychological support from their partners and will have easy access to safe abortion services. A study in Nepal found that unmarried women often face different barriers to receiving safe abortion services relative to their married counterparts. ^33^ The findings of this study further revealed that women in urban areas were less likely to engage in unsafe abortion methods compared to their rural counterparts. Similar results were found by earlier studies in Ghana ^20 25^ and Mozambique. ^35^ The possible explanation to this is the lack of access to safe abortion services among women in the rural areas. This compels them to resort to the use of unsafe methods of abortion. Further, the conservative cultural belief system and perception about abortion often compels rural women to secretly practice abortion which is often unsafe.

Knowledge and awareness on the law governing abortion practice can influence women with unplanned pregnancies to have access to safe abortion methods and use safe abortion methods. ^25 51^ In this study, respondents’ knowledge within the decade (2007-2017) on the legal status of abortion in Ghana have increased but this has not reflected in the reduction of the number of women who use unsafe abortion method as the proportion who used safe abortion methods have rather decreased. A study by Coast et al ^16^ provided some explanation to this phenomenon. They argue that women’s knowledge on the legal status of abortion may be negatively influenced by the wider social norms, cultural belief system, religious regulations and conventions surrounding abortion might coerced them to go for unsafe methods of abortion which usually does not occur in a public space. Further, women knowledge on the legality of abortion might prevent them from seeking safer abortion services because of financial support especially from the partners since safe methods are relatively expensive to unsafe abortion practices.^52 53^ Also, studies have found that knowledge on legal status of abortion is low among some health professionals ^54^ and this negatively influences service provider attitudes towards privacy and stigmatising provider behaviours. ^55^ Women will rather prefer to practice unsafe abortions in order to avoid being victim of stigmatization, labelling and emotional trauma.

There are some limitations to this study that must be recognised. First and foremost, not all factors that are associated with choice of abortion methods have been explored. Further, this study used a secondary cross-sectional data which limited us from asking further question to get deeper understanding of the phenomena. Finally, the use of cross-sectional data involved a recall of past events over a five-year period. This means there are possibilities of recall bias and under-reporting of abortion-related activities. Nonetheless, we use a nationally representative data and robust statistical methods and analyses to examine the subject matter under investigation. It can therefore be generalised to women in Ghana.

## Conclusion

Abortion is a sensitive issue in the Africa region because of the legal, cultural, social and moral connotations that surroundings it and has implication on the fertility, morbidity and mortality trends in the African region. In this study, increasing knowledge of legal status on abortion among women interestingly correspond with an increased in the use of unsafe abortion methods in Ghana. It is recommended that social norms, cultural believes and misconceptions that inhibit women access to safe abortion services should be relaxed to enable them to have easy access to these services in the African region. This is because if a woman knows the abortion laws of Ghana prohibit abortion unless under specific circumstances (rape, incest, mother/foetus health at risk), but the social, cultural and religious norms does not support abortion, she might practice the unsafe method which is mostly done secretly. The liberalisation of social, cultural and religious regulations especially in the rural areas might help women with unplanned pregnancy to opt for safer abortion care.

## Data Availability

All data relevant to the study uploaded as supplementary information. Data is however openly available and can be accessed via https://dhsprogram.com/

## Contributorship

DK conceptualised and design the study. DK and DYA analysed and interpreted the data. DK also drafted the entire manuscript. AKM, MD and MI reviewed and revised the manuscript. All authors approved the final version of the article.

## Patient and public involvement

No patient involved in the design, or conduct, or reporting of this research

## Patient consent for publication

Not required

## Appendices

## Appendix 1

## Appendix 2

## Appendix 3

## Appendix 4

## Reference

1. Sedgh G, Bearak J, Singh S, Bankole A, Popinchalk A, Ganatra B, Rossier C, Gerdts C, Tunçalp Ö, Johnson Jr BR, Johnston HB. Abortion incidence between 1990 and 2014: global, regional, and subregional levels and trends. The Lancet. 2016 Jul 16;388(10041):258–67.

2. Ganatra B, Gerdts C, Rossier C, Johnson Jr BR, Tunçalp Ö, Assifi A, Sedgh G, Singh S, Bankole A, Popinchalk A, Bearak J. Global, regional, and subregional classification of abortions by safety, 2010–14: estimates from a Bayesian hierarchical model. The Lancet. 2017 Nov 25;390(10110):2372–81.

3. Singh S, Remez L, Sedgh G, Kwok L, Onda T. Abortion worldwide 2017: uneven Progress and unequal AccessAbortion worldwide 2017: uneven Progress and unequal Access.

4. DePiñeres T, Raifman S, Mora M, Villarreal C, Foster DG, Gerdts C. ‘I felt the world crash down on me’: Women’s experiences being denied legal abortion in Colombia. Reproductive health. 2017 Dec;14(1):1–9.

5. Say L, Chou D, Gemmill A, Tunçalp Ö, Moller AB, Daniels J, Gülmezoglu AM, Temmerman M, Alkema L. Global causes of maternal death: a WHO systematic analysis. The Lancet Global Health. 2014 Jun 1;2(6):e323–33.

6. Morhee RA, Morhee ES. Overview of the law and availability of abortion services in Ghana. Ghana medical journal. 2006;40(3).

7. Sundaram A, Juarez F, Bankole A, Singh S. Factors associated with abortion]seeking and obtaining a safe abortion in Ghana. Studies in family planning. 2012 Dec;43(4):273–86.

8. Ghana. Statistical Service, Macro Systems. Institute for Resource Development. Demographic, Health Surveys. Ghana demographic and health survey. Ghana Statistical Service; 2008.

9. Morhe ES, Morhe RA, Danso KA. Attitudes of doctors toward establishing safe abortion units in Ghana. International Journal of Gynecology & Obstetrics. 2007 Jul 1;98(1):70–4.

10. Johnson Jr BR, Mishra V, Lavelanet AF, Khosla R, Ganatra B. A global database of abortion laws, policies, health standards and guidelines. Bulletin of the World Health Organization. 2017 Jul 1;95(7):542.

11. List GE. Ghana National Drugs Programme (GNDP) Ministry of Health (MOH), Republic of Ghana. Ghana Essential Medicines List.

12. Aladago DA, Boakye-Yiadom A, Asaarik MJ, Aryee PA. THE CONSEQUENCES OF ABORTION RESTRICTIONS FOR ADOLESCENTS’HEALTHCARE IN GHANA: THE INFLUENCE OF GHANA’S ABORTION LAW ON ACCESS TO SAFE ABORTION SERVICES. UDS International Journal of Development. 2019;6(1):1–9.

13. Biggs MA, Gould H, Foster DG. Understanding why women seek abortions in the US. BMC women’s health. 2013 Dec 1;13(1):29.

14. McLemore MR, Kools S, Levi AJ. Calculus formation: Nurses’ decision]making in abortion]related care. Research in nursing & health. 2015 Jun;38(3):222–31.

15. Eisenberg DL, Leslie VC. Threats to reproductive health care: time for obstetrician-gynecologists to get involved. American journal of obstetrics and gynecology. 2017 Mar 1;216(3):256–e1.

16. Coast E, Norris AH, Moore AM, Freeman E. Trajectories of women’s abortion-related care: A conceptual framework. Social Science & Medicine. 2018 Mar 1;200:199–210.

17. Darroch JE, Singh S, Weissman E. Adding it up: the costs and benefits of investing in sexual and reproductive health 2014—estimation methodology. Appendix B: estimating sexual and reproductive health program and systems costs. New York: Guttmacher Institute. 2016.

18. Konney TO, Danso KA, Odoi AT, Opare-Addo HS, Morhe ES. Attitude of women with abortion-related complications toward provision of safe abortion services in Ghana. Journal of Women’s Health. 2009 Nov 1;18(11):1863–6.

19. Owoo NS, Lambon-Quayefio MP, Onuoha N. Abortion experience and self-efficacy: exploring socioeconomic profiles of GHANAIAN women. Reproductive health. 2019 Dec 1;16(1):117.

20. Mundigo AI. Determinants of unsafe induced abortion in developing countries. Unsafe Abortion. 2006;51.

21. Payne CM, Debbink MP, Steele EA, Buck CT, Martin LA, Hassinger JA, Harris LH. Why women are dying from unsafe abortion: narratives of Ghanaian abortion providers. African Journal of Reproductive Health. 2013 Jul 4;17(2):118–28.

22. Klutsey EE, Ankomah A. Factors associated with induced abortion at selected hospitals in the Volta Region, Ghana. International journal of women’s health. 2014;6:809.

23. Sousa A, Lozano R, Gakidou E. Exploring the determinants of unsafe abortion: improving the evidence base in Mexico. Health Policy and Planning. 2010 Jul 1;25(4):300–10.

24. Boah M, Bordotsiah S, Kuurdong S. Predictors of unsafe induced abortion among women in Ghana. Journal of pregnancy. 2019 Feb 3;2019.

25. Reiger ST, Dako-Gyeke P, Ngo TD, Eva G, Gobah L, Blanchard K, Grindlay K. Abortion knowledge and experiences among young women and men in Accra, Ghana. Gates Open Research. 2019 May 30;3(1478):1478.

26. Gbagbo FY. Women’s Prior Knowledge of The Abortion Law and Decision-Making on Choice of Place for Abortion Services in Accra, Ghana. Mathews J Gynecol Obstet. 2019;4(1):17.

27. Cresswell JA, Schroeder R, Dennis M, Owolabi O, Vwalika B, Musheke M, Campbell O, Filippi V. Women’s knowledge and attitudes surrounding abortion in Zambia: a cross-sectional survey across three provinces. BMJ open. 2016 Mar 1;6(3).

28. Assifi AR, Berger B, Tunçalp Ö, Khosla R, Ganatra B. Women’s awareness and knowledge of abortion laws: A systematic review. PloS one. 2016 Mar 24;11(3):e0152224.

29. Schuster S. Abortion in the moral world of the Cameroon grassfields. Reproductive Health Matters. 2005 Jan 1;13(26):130–8.

30. Ngowa JD, Neng HT, Domgue JF, Nsahlai CJ, Kasia JM. Voluntary induced abortion in Cameroon: prevalence, reasons, and complications. Open Journal of Obstetrics and Gynecology. 2015;5(09):475.

31. Bongfen MC, Abanem EE. Abortion practices among women in Buea: a socio-legal investigation. The Pan African Medical Journal. 2019;32.

32. Andersen KL, Khanal RC, Teixeira A, Neupane S, Sharma S, Acre VN, Gallo MF. Marital status and abortion among young women in Rupandehi, Nepal. BMC women’s health. 2015 Dec 1;15(1):17.

33. Atakro CA, Addo SB, Aboagye JS, Menlah A, Garti I, Amoa-Gyarteng KG, Sarpong T, Adatara P, Kumah KJ, Asare BB, Mensah AK. Contributing factors to unsafe abortion practices among women of reproductive age at selected district hospitals in the Ashanti region of Ghana. BMC women’s health. 2019 Dec 1;19(1):60.

34. Dickson KS, Adde KS, Ahinkorah BO. Socio–economic determinants of abortion among women in Mozambique and Ghana: evidence from demographic and health survey. Archives of Public Health. 2018 Dec;76(1):37.

35. Agrawal S. Determinants of Induced Abortion and Its Consequences on Women’s Reproductive Health: Findings from India’s Natinal Family Health Surveys. Macro International; 2008.

36. Cook PJ, Parnell AM, Moore MJ, Pagnini D. The effects of short-term variation in abortion funding on pregnancy outcomes. Journal of Health Economics. 1999 Apr 1;18(2):241–57.

37. Rahman A, Katzive L, Henshaw SK. A global review of laws on induced abortion, 1985-1997. International Family Planning Perspectives. 1998 Jun 1:56–64.

38. Schwelb E. The Republican Constitution of Ghana. The American Journal of Comparative Law. 1960 Oct 1:634–56.

39. Sundaram A, Juarez F, Bankole A, Singh S. Factors associated with abortion]seeking and obtaining a safe abortion in Ghana. Studies in family planning. 2012 Dec;43(4):273–86.

40. Leone T, Coast E, Parmar D, Vwalika B. The individual level cost of pregnancy termination in Zambia: a comparison of safe and unsafe abortion. Health policy and planning. 2016 Sep 1;31(7):825–33.

41. Mote CV, Otupiri E, Hindin MJ. Factors associated with induced abortion among women in Hohoe, Ghana. African journal of reproductive health. 2010;14(4).

42. Shah IH, Åhman E. Unsafe abortion differentials in 2008 by age and developing country region: high burden among young women. Reproductive health matters. 2012 Jan 1;20(39):169–73.

43. Dehlendorf C, Weitz T. Access to abortion services: a neglected health disparity. Journal of health care for the poor and underserved. 2011;22(2):415–21.

44. Jones RK, Jerman J. How far did US women travel for abortion services in 2008?. Journal of women’s health. 2013 Aug 1;22(8):706–13.

45. Ostrach B, Cheyney M. Navigating social and institutional obstacles: low-income women seeking abortion. Qualitative health research. 2014 Jul;24(7):1006–17.

46. Ghana Statistical Service, Ghana Health Service, and ICFGhana Maternal Health Survey, Accra, 2007. Ghana,

47. Ghana Statistical Service, Ghana Health Service, and ICF, Ghana Maternal Health Survey Accra, Ghana 2018.

48. Wodajo LT, Mengesha ST, Beyen TK. Unsafe abortion and associated factors among women in reproductive age group in Arsi Zone, Central Ethiopia. International Journal of Nursing and Midwifery. 2017 Oct 31;9(10):121–8.

49. Yogi A, Prakash KC, Neupane S. Prevalence and factors associated with abortion and unsafe abortion in Nepal: a nationwide cross-sectional study. BMC pregnancy and childbirth. 2018 Dec 1;18(1):376.

50. Marlow HM, Wamugi S, Yegon E, Fetters T, Wanaswa L, Msipa-Ndebele S. Women’s perceptions about abortion in their communities: perspectives from western Kenya. Reproductive Health Matters. 2014 May 1;22(43):149–58.

51. Nyanzi S, Nyanzi B, Bessie K. “Abortion? That’s for women!” Narratives and experiences of commercial motorbike riders in south-western Uganda. African journal of reproductive health. 2005 Apr 1:142-61.

52. Schwandt HM, Creanga AA, Adanu RM, Danso KA, Agbenyega T, Hindin MJ. Pathways to unsafe abortion in Ghana: the role of male partners, women and health care providers. Contraception. 2013 Oct 1;88(4):509–17.

53. Calvès AE. Abortion risk and decisionmaking among young people in urban Cameroon. Studies in Family Planning. 2002 Sep;33(3):249–60.

54. Labandera A, Gorgoroso M, Briozzo L. Implementation of the risk and harm reduction strategy against unsafe abortion in Uruguay: From a university hospital to the entire country. International Journal of Gynecology & Obstetrics. 2016 Aug;134(S1):S7–11.

55. DaVanzo J, Rahman M. Pregnancy termination in Matlab, Bangladesh: trends and correlates of use of safer and less-safe methods. International perspectives on sexual and reproductive health. 2014 Sep 1;40(3):119–26.

56. Thapa S, Neupane S, Basnett I, Read E. Women having abortion in urban Nepal: 2005 and 2010 compared. Kathmandu University medical journal. 2012;10(3):8–13.

57. Pallikadavath S, Stones RW. Maternal and social factors associated with abortion in India: a population-based study. International Family Planning Perspectives. 2006 Sep 1:120–5.

